# Comparison of a Commercially Available Prostate Segmentation Application to Traditional Prolate and Biproximate Ellipsoid Methods for Prostate Volume Measurement

**DOI:** 10.1101/2020.10.21.20216374

**Authors:** Neil F. Wasserman, Benjamin Spilseth

## Abstract

**Rationale and Objectives:** To compare the a commercially available automatic and manually adjusted segmentation software program (DynaCAD ^®^) to two ellipsoid volume methods using T2-weighted magnetic resonance imaging (MRI).

**Material and Methods:** This is a retrospective IRB-approved study of 146 patients randomly selected from 1600 consecutive men referred for T2-weighted MRI. All measurements were performed by a single expert senior radiologist. Total prostate volume was calculated using automatic DynaCAD ^®^ software (RCAD), manually adjusted DynaCAD ^®^ (ACAD), traditional ellipsoid method (TE) and a new alternative biproximate ellipsoid method (BE). Results were assessed with ANOVA and linear regression.

**Results:** Mean volumes for RCAD, ACAD, BE and TE were 61.5, 58.4, 56, and 53.2 respectively. ANOVA showed no difference of the means (p> 0.05.) Linear regression showed a coefficient of determination (r ^2^) between ACAD and TE of 0.92 and between ACAD and BE of 0.90. Using the planigraphic-based segmented ACAD as the “gold’ standard, RCAD overestimated volume by 5%. TE and BE underestimated prostatic volume by 4% and 9% respectively. ACAD processing time was 4.5 to 9.5 minutes (mean=6.6 min.) compared to 1.5 to 3.0 minutes (mean=2.3 min.) for prolate ellipsoid methods.

**Conclusion:** Manually adjusted MRI T2-weighted segmentation is likely the most accurate measure of total prostate volume. DynaCAD appears to fulfill that function, but manual adjustment of automatic misregistration of boundaries is necessary. ACAD and RCAD are best applied to research use. Ellipsoid methods are faster, more convenient, nearly as accurate and more practical for clinical use.

## Introduction

Various imaging modalities and methods have been used in the past for calculation of prostatic volume in order to stratify clinical decision–making for medical, surgical or minimally invasive treatment of patients with prostate cancer [1] and benign prostatic hyperplasia (BPH) [2]. Christie, DRH, et al [3] extensively reviewed and compared the use of ultrasound, computed tomography and magnetic resonance imaging (MRI) in volume measurement of the prostate. Littrup, et al [1] compared transrectal ultrasound (TRUS) planigraphy with various formula-based ellipsoid methods under the assumption that planigraphy was the imaging standard to which ellipsoid techniques should be compared. They demonstrated that the ellipsoid formula compared favorably to ultrasound planigraphy. Many studies appeared in the ultrasound literature applying ellipsoid formulae due to its speed and ease of use in the clinical setting [1, 4, 5]. The use of the ellipsoid formula later migrated to MRI as a clinical method for estimating total prostate volume [6-8] until more recent research on MRI-based planigraphy entered the medical literature [9, 10]. There are few commercially available prostate software programs based on MRI planigraphy.

More recently, the term *segmentation* has replaced planigraphy for computer-generated boundary-finding algorithms used to separate selected volumes of interest from surrounding tissues. At present, artificial intelligent applications for prostate volume segmentation remains largely a research enterprise.

One popular commercially available product for MRI-based segmentation is DynaCAD^®^ (Invivo Corporation, Gainesville, FL). However, this software lacks convincing proof of efficacy in the field. There is only one study in the medical literature where it was used [10]. The manufacturer does not disclose its theoretical base or algorithm because it is “proprietary”. This product lacks published Dice similarity coefficient or other standard measures. Additionally, the developer reports no in-house studies of accuracy and precision, nor are they aware of testing or publishing of its use in the field. (Personal communication with Invivo technical support engineer (2018,2020).

The purpose of this report is to compare DynaCAD^®^ performance to the traditional ellipsoid (TE) and recently published biproximate ellipsoid (BE) methods. The latter was developed for greater inter-observer consistency (precision) using MRI [11]. Can we rely on unadjusted (fully automatic) or manually adjusted DynaCAD^®^ for measurement of prostatic volume? If so, what are its limitations? And, finally, is DynaCAD^®^ or any other automated or semiautomated segmentation software practical at this time for clinical use?

## Materials and Methods

### Patient Population and Selection

This is a retrospective IRB and Ethics Committee-approved study # 2871 approved study of 1600 consecutive patients referred for prostate MRI because of elevated serum prostate specific antigen (PSA) levels, abnormal digital rectal examination (DRE) or both from the years January 2016 through December 2017. From this initial study cohort, those with previous history of prostate surgery, minimally invasive prostate therapy, or radiation were excluded. Of the remaining 1447 cases, final study cohort was obtained by randomly selecting every tenth consecutive patient resulting in 144 cases.

### MRI Techniques

Prostate MRI was performed using 3T for all patients (Magnetom Skyra or TrioTim; Siemens Healthcare; Erlangen Germany). The majority were performed using a pelvic phase array protocol without endorectal coil (ERC), and less that 10% were performed with Endorectal coil (Medrad, NJ, USA). The exact imaging parameters evolved during the study period, and at all times PI-RADS compliant technique was utilized. A typical protocol during the study period included T2W fast spin echo (FSE) imaging in the axial plane (TR/TE 3700/111 msec; NEX 3; 3mm slice thickness; no interslice gap; flip angle 160 deg; FOV 140 mm, matrix 320 × 256) and coronal plane (TR/TE 4030/100 msec; NEX 2; 3 mm slice thickness; no interslice gap; flip angle 122 deg; FOV 180 mm; matrix 320 × 256). T2W 3D SPACE images were obtained (TR/TE 1400/101 msec; flip angle 135 deg; 256 × 256 × 205 matrix, 180 mm FOV). T2W 3D SPACE images were reconstructed at 1 mm in the axial plane and also at 3mm in the, axial, sagittal, and coronal planes. Additionally, dynamic contrast enhanced T1-weighted (3 mm slice thickness; TR/TE 4.9/1.8 msec; 224 × 156 matrix, 250 mm FOV, temporal resolution <10 sec), and diffusion-weighted images at b values of 50, 800, and 2000 s/mm3 were performed. Endorectal coils were used in 2 patients.

### Image Analysis

All measurements were performed by an expert senior radiologist with over 40-years-experience with TRUS and MRI prostate volumetrics. The images were reviewed on high-resolution monitors Philips Intellispace portal (Koninklijke Philips N.V.). Measurements for TE and BE were done in alternating order using standard PAC tools (Fig. 1). The ellipsoid formula applied was:

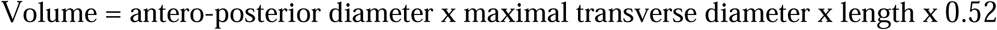

**Fig. 1.**
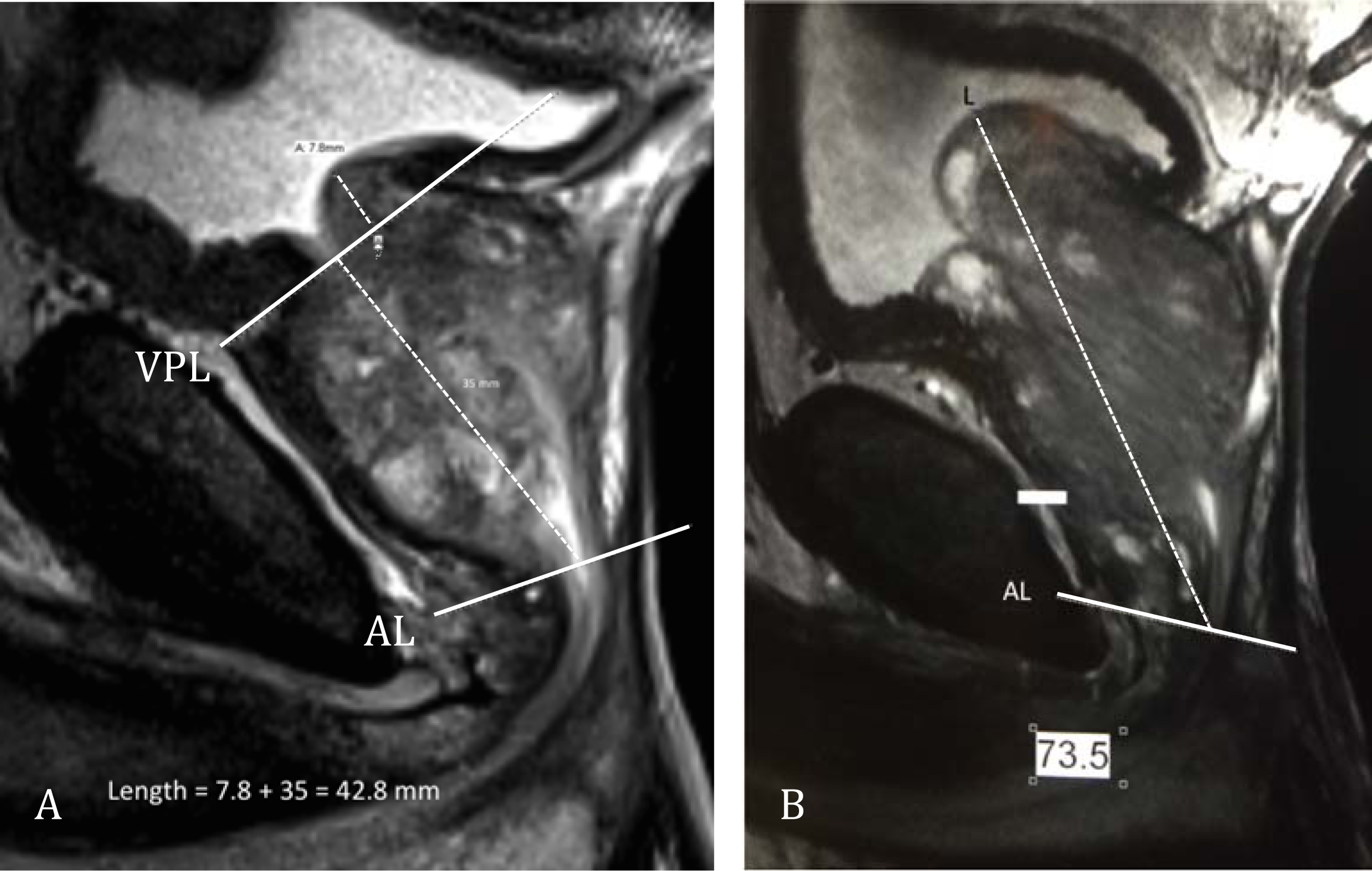
MRI T2-weighted mid-sagittal views of two different prostates. A. Demonstrates biproximate method of length measurement and B illustrates traditional length measurement from apex to most proximal tissue. AL=apical line, VPL=vesical-prostatic line. (Details can be found in reference 10)

The entire image sequence was then migrated using DICOM to DynaCAD^®^ software. The automatic volumes were recorded. Manual adjustments of boundaries were then performed at all axial 3mm intervals from prostate base to apex resulting in adjusted values that were recorded for comparison. The final matrix of volumetrics included raw unadjusted DynaCAD, (RCAD) manually adjusted DynaCAD (ACAD), traditional ellipsoid (TE) and biproximate ellipsoid (BE) data. Detailed description of the biproximate technique has been described [11]. Determination of whether a method over-or underestimated prostatic volume compared to the ACAD was calculated by dividing the difference in the 2 means by the larger of the 2 means and multiplying by 100 to derive the percent of change.

### Statistics

Means of the four methods were compared for significant differences using ANOVA as the data were normally distributed. The criterion for significant difference was p<0.05. Direct comparisons of subjects’ prostate volumes utilizing each method were analyzed using Pearson product correlation coefficient and linear logistic regression. Bland Altman plots were calculated to appraise agreement between methods. All statistics were calculated using QI Macro Statistics^®^ (KnowWare International, Inc., Denver CO, USA).

## Results

Median age of the final cohort was 61-years, (range, 27-84). Median PSA =7.1 ng/mL (range, 0.58-145.4). Mean volumes RCAD, manually ACAD, biproximate ellipsoid and traditional ellipsoid are shown in Table 1. ANOVA showed no significant difference in the means (p=0.41). Respective median values of 46.6, 43.7, 41.5, 43.5 are also shown in Figure 2 box plot.

**Table 1.** Central Values for 4 Methods of Total Prostate Volume Measurement in cc (n=144)*

| Method | Raw CAD | Adj. CAD | Biproximate | Traditional |
| --- | --- | --- | --- | --- |
| Mean | 61.5 | 58.4 | 53.2 | 56.0 |
| CI | 54.3-68.8 | 51.1-65.8 | 46.5-59.9 | 49.1-63.1 |
| Median | 46.6 | 43.7 | 41.5 | 53.5 |

\*No difference in means at $p>0.05$

**Fig. 2.**
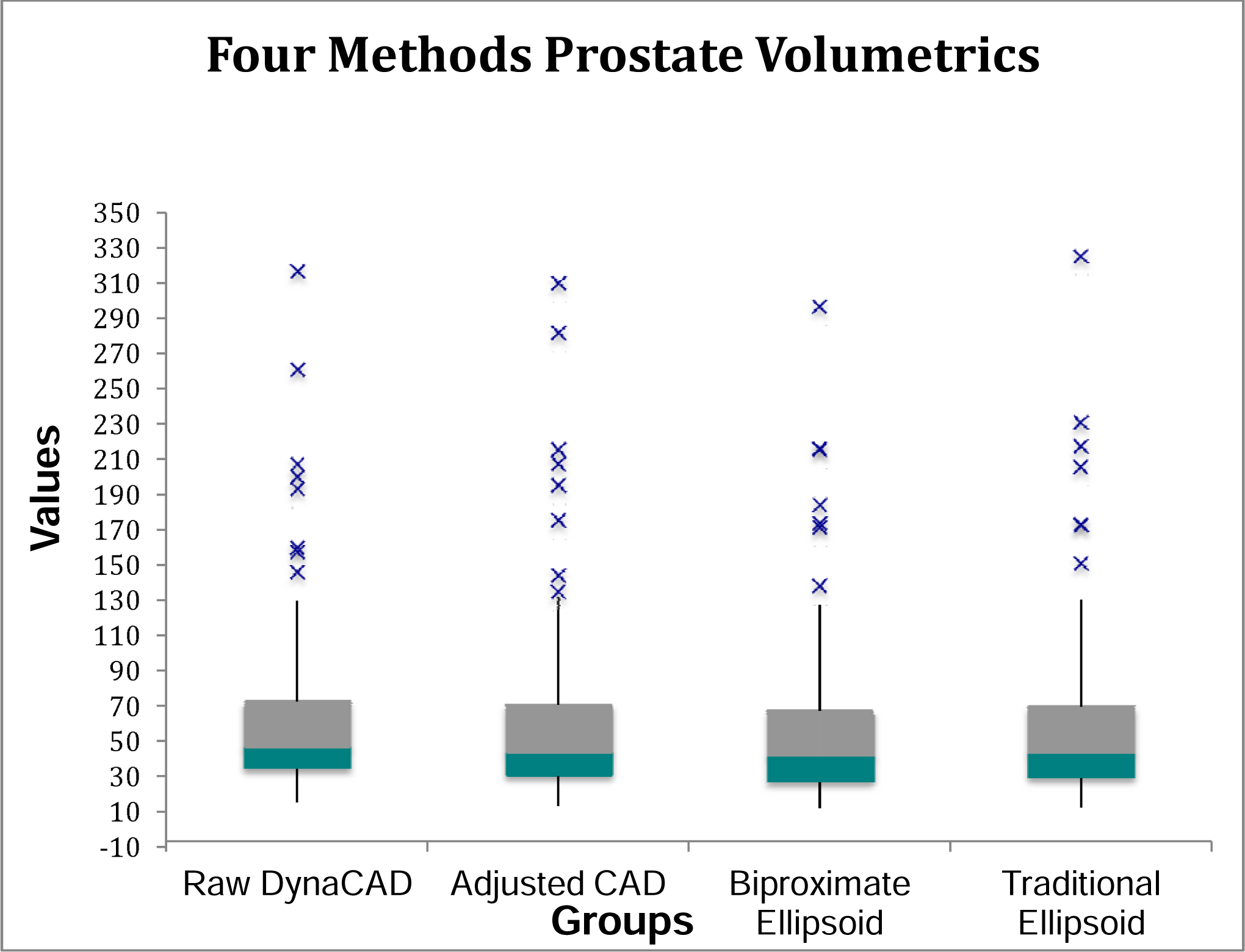
Box chart showing distribution of measurements for Raw (automatic) CAD prostate volume, Adjusted CAD, Biproximate Ellipsoid, and Traditional Ellipsoid groups. Interface within boxes represent median values

Linear regression of ACAD versus RCAD resulted in an r ^2^ of 0.93 as shown in Fig. 3A. Linear regression of ACAD versus traditional ellipsoid resulted in an r^2^ of 0.921 (Fig. 3B) while linear regression of the ACAD versus BE method showed an r^2^ of 0.90 (Fig. 3C). TE and BE showed an r^2^ of 0.99 in our study (Fig. 3D). Bland Altman plots are shown in Fig. 4A and B.

**Fig. 3.**
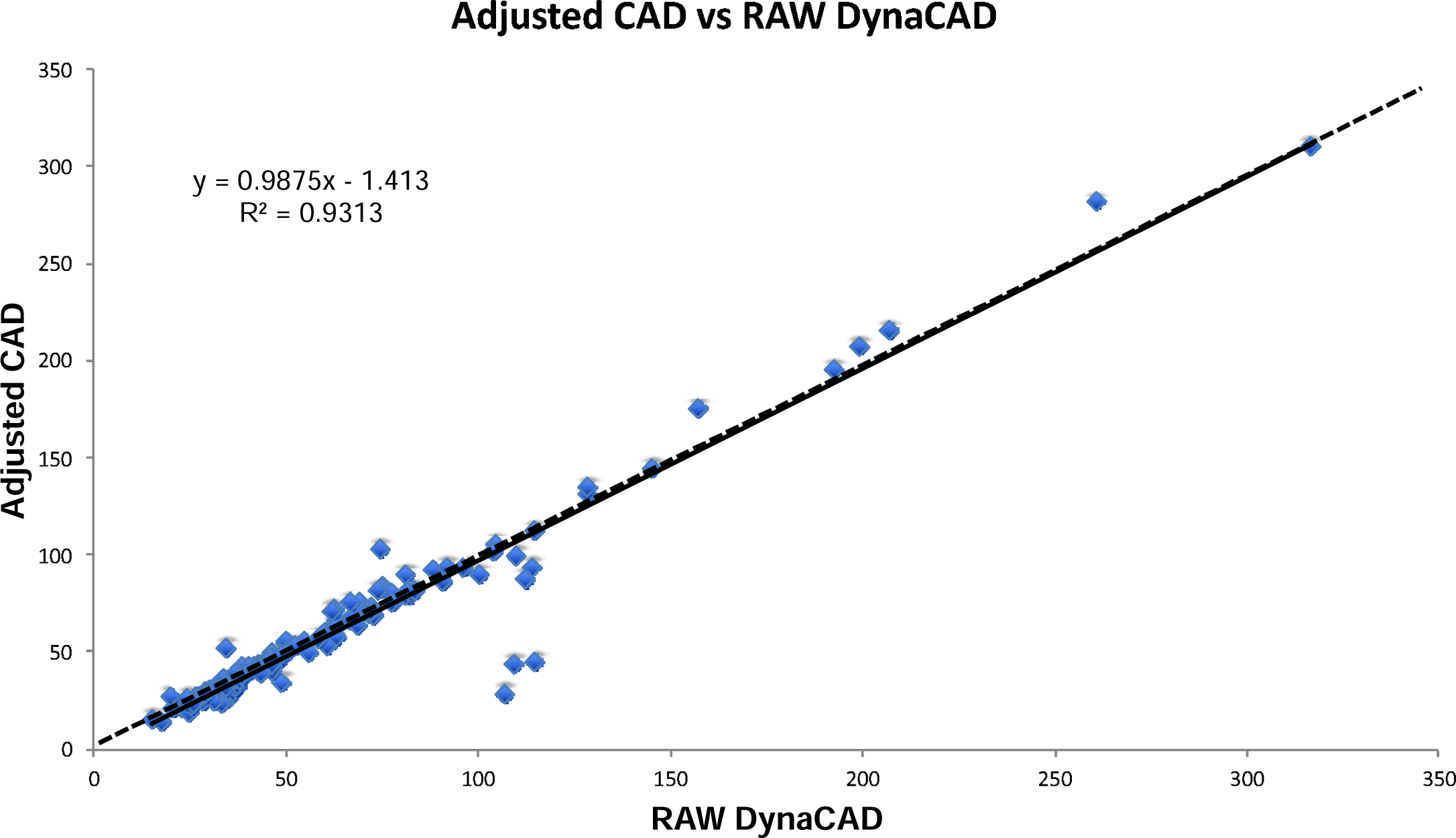
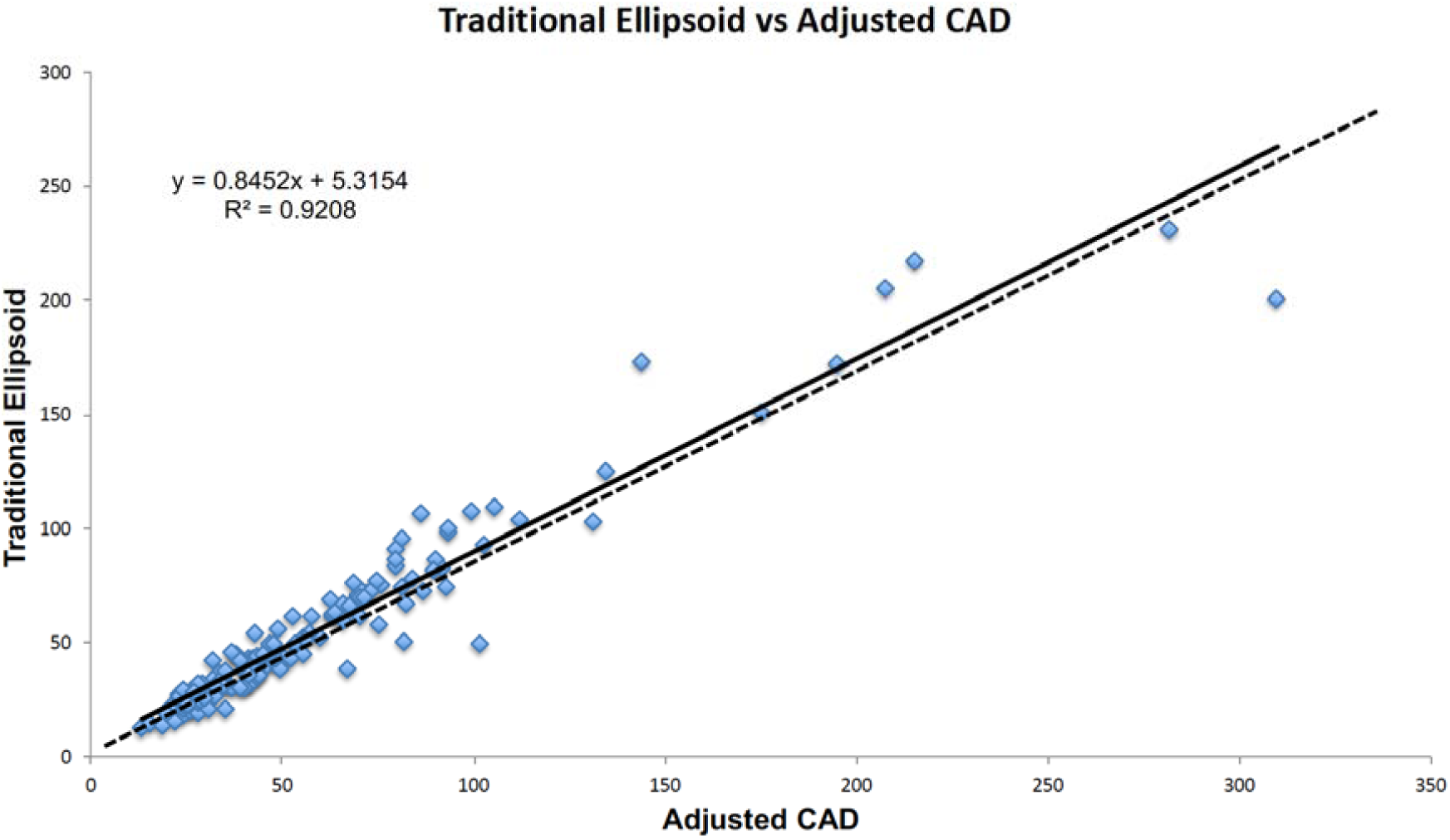
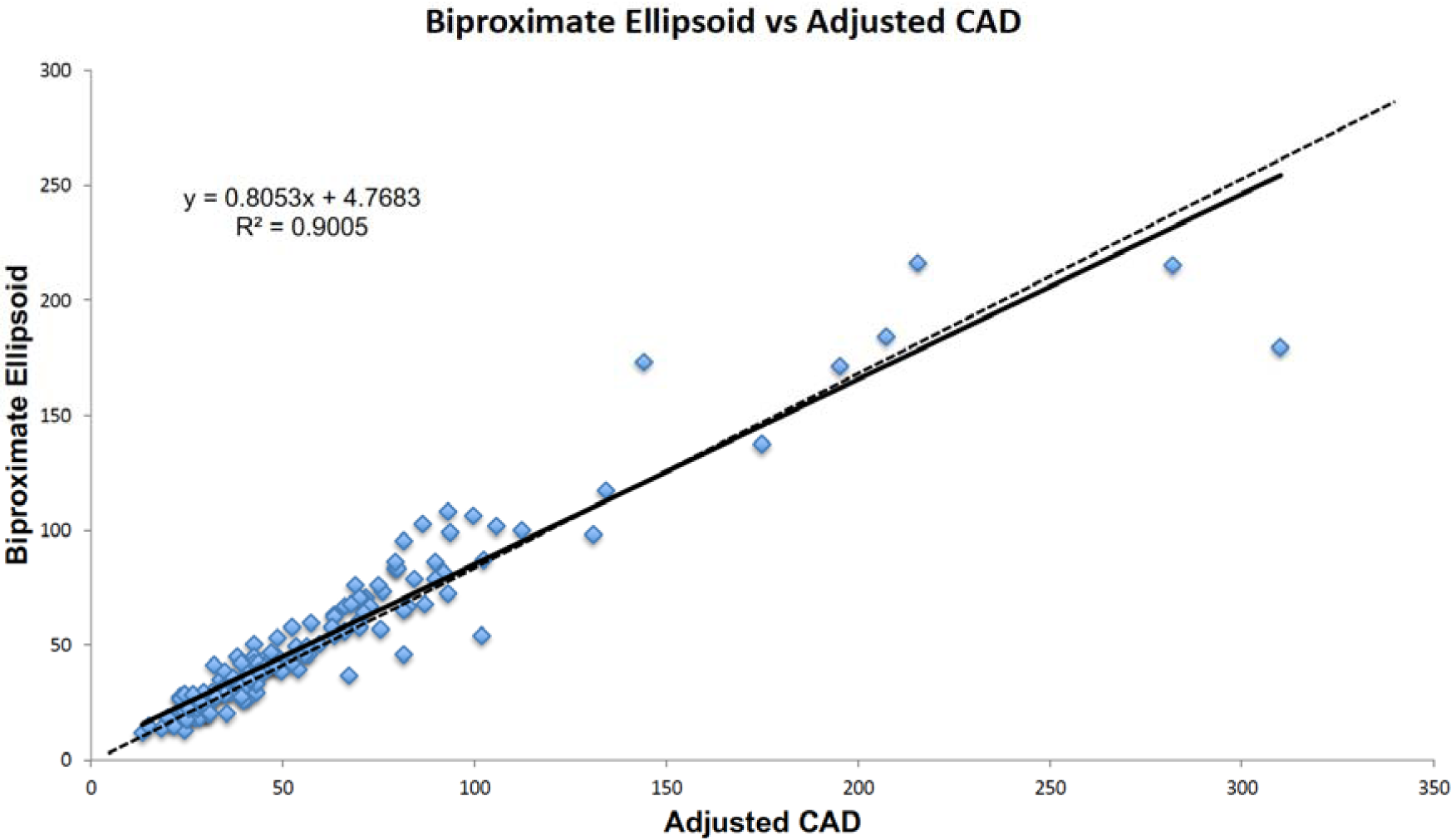
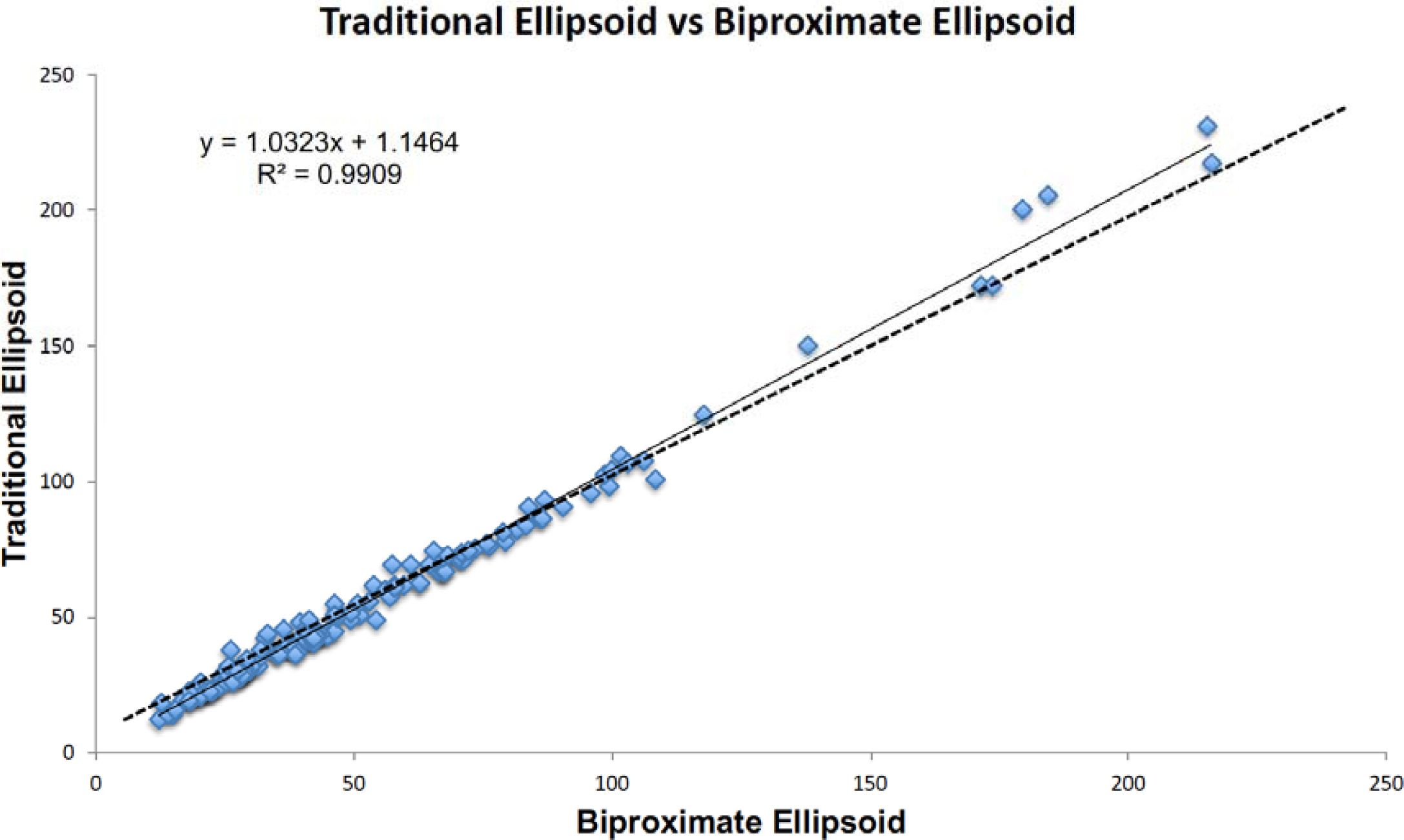
Linear regression lines comparing manually Adjusted CAD with Raw (Unadjusted) CAD A, ACAD with Traditional Ellipsoid volumes B, Biproximate vs Traditional Ellipsoid volumes C, and Traditional vs Biproximate Ellipsoid volumes D. Dashed line is line of equality (Ref. 11)

**Fig. 4.**
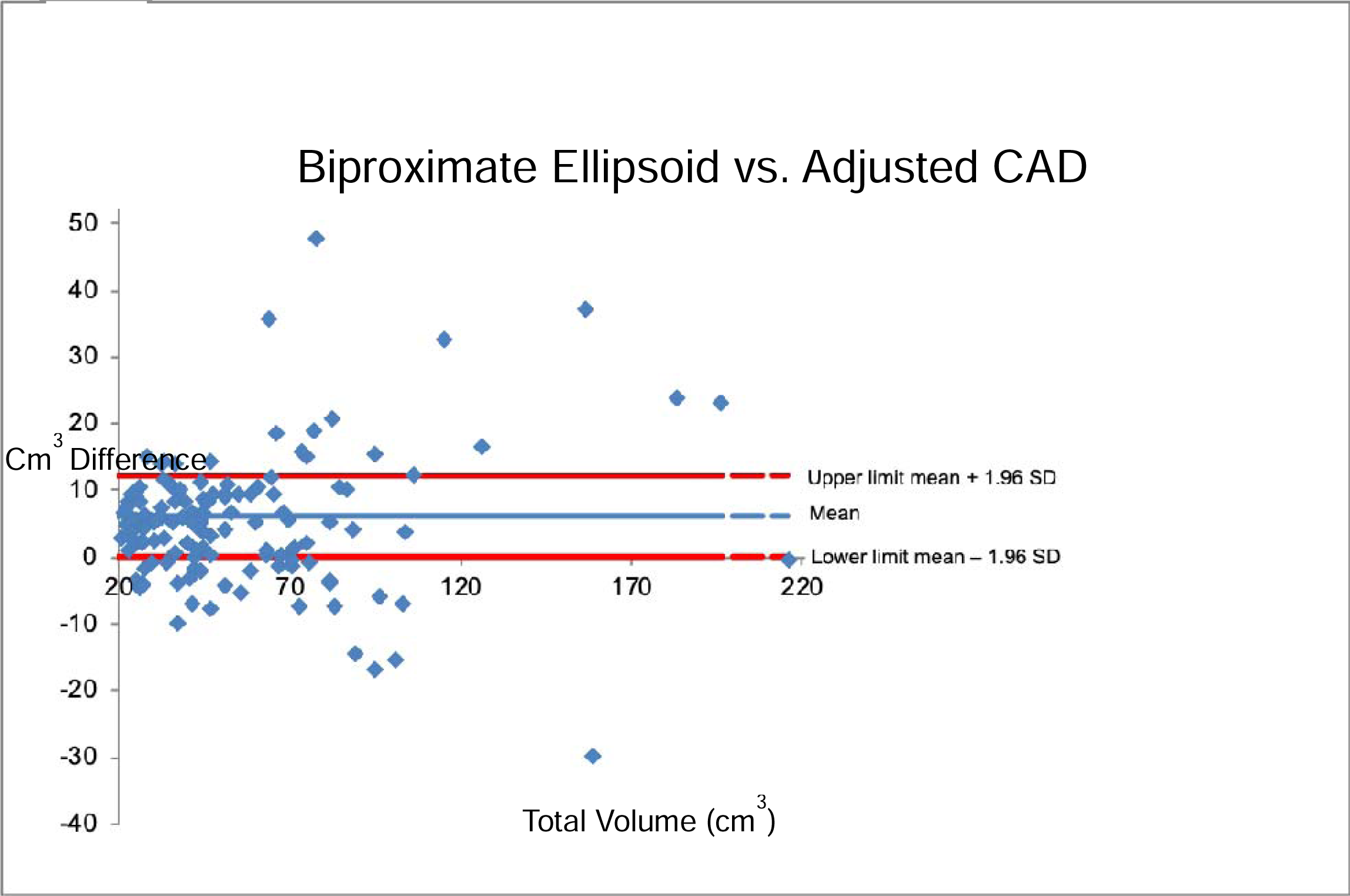
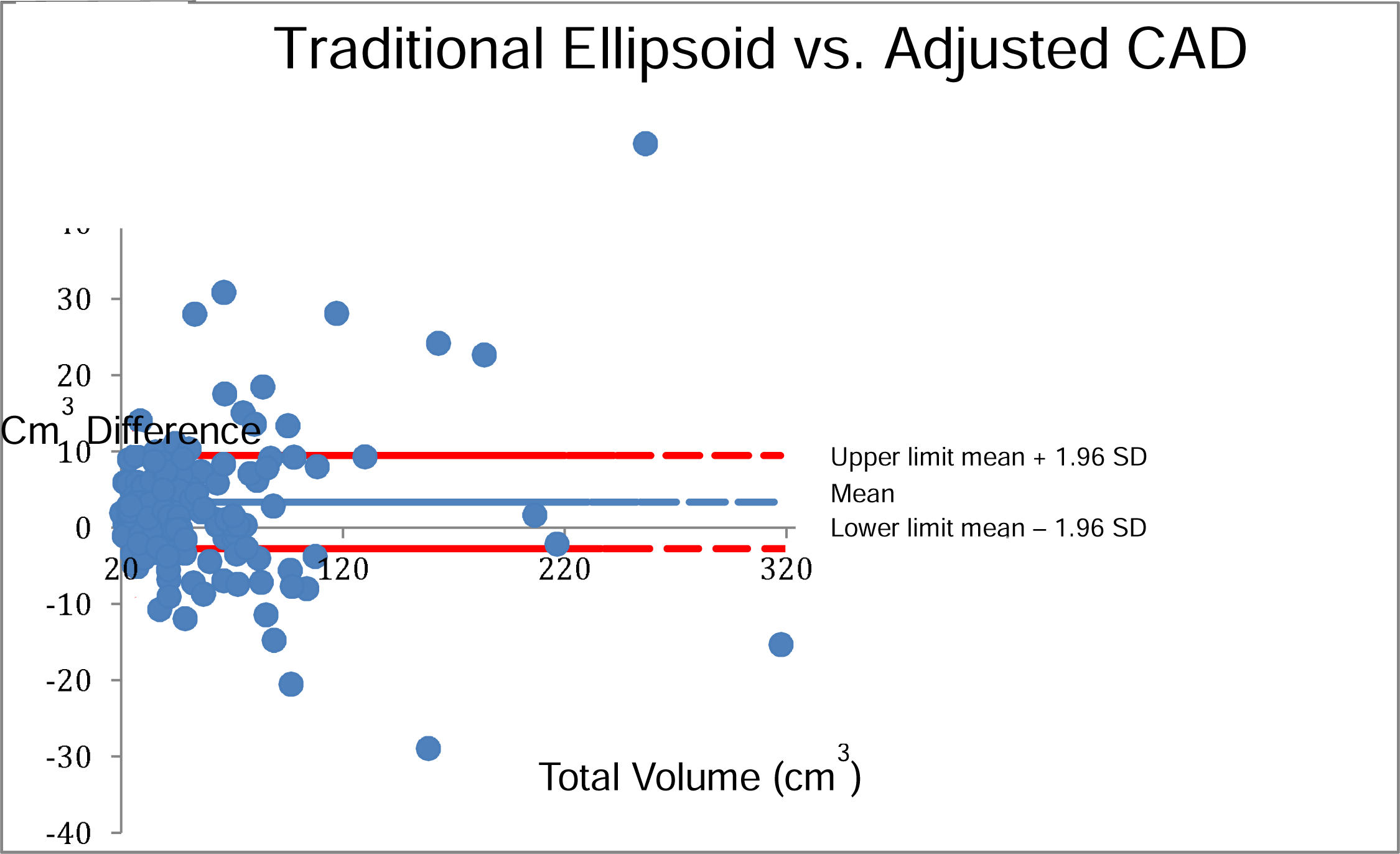
Bland Altman Plots for Biproximate Ellipsoid (A) and Traditional Ellipsoid methods (B) for measuring total prostate volume. Each dot/diamond represents the difference in cm^3^ of two measurements on the same patient. Two-thirds of the measures fall within the 95% limits of agreement for Biproximate and 75% for Traditional. SD=standard deviation

We took ACAD as the “gold standard” with which to compare RCAD and ellipsoid models in this study. RCAD, overestimated total prostatic volume by 5% compared with ACAD. The TE technique underestimated total prostatic volume by 4% compared with ACAD, while the BE method underestimated volume by 9%. Thus, BE underestimated traditional ellipsoid by 5.%.

Not counting transfer time from PACS to DynaCAD^®^ the time required for manual segmentation was quite variable. This examiner was very deliberate resulting in total times between 4.5 and 9.5 minutes to adjust boundaries at all levels and in at least two views. Some fully automated patients needed adjustment at fewer levels. Ellipsoid methods took between 1.5 and 3.0 minutes. Although of interest, more detailed statistical analysis of observer times was not performed in this investigation because one individual’s experience wasn’t thought to be translatable to the real world experience.

## Discussion

Our results with MRI compare favorably with that of Littrup using TRUP. MRI showed r^2^ of 0.921 for TE and 0.90 for BE versus ACAD compared with r^2^ of 0.894 and 0.856 with TRUS planigraphy [1]. The results demonstrate that in comparing the two ellipsoid techniques to ACAD there is not only high correlation but also minimal bias as evidenced by the close proximity of the slope to the line of equality (dashed line in figures). The line of equality is derived from theoretic measurements of a randomly selected cohort resulting in almost perfect agreement [12]. If the regression lines were well above or below the equality line, it would indicate a consistent error of measurement due to bias in the measurements of the observer [12]. One advantage of a single observer is that it minimizes the likelihood of bias since the same examiner is measuring the cohort with all volumetric techniques.

It has been pointed out that high correlation between 2 measuring tests does not necessarily equate to high “agreement” [12]. Many believe that a Bland Altman plot of the difference between two measurements of the same individual sample provides a better visual display for understanding the limits of agreement [12,13]. Bland Altman plots are displayed for this study in Figs. 4A and B). Since the majority of the plotted differences fall between the 95% limits (especially at lower prostate volumes), and considering the clinical setting, agreement is considered adequate for clinical purposes.

TE and BE methods underestimated prostate volume using manually ACAD as the standard. This most likely is due to our methods wherein we chose to measure dimensions from the inner boundary of the external prostatic capsule while DynaCAD^®^ measures from the outer boundary. The thickness of the external capsule at the measurement level is 1-1.5 mm which when summed at each level and added to the axial measurement accounts for much of this discrepancy.

ACAD was selected over RCAD as the “gold standard” because it likely more accurately approximates true planigraphy. This has been generally accepted as corresponding best to prostate specimen weight because of more precise inclusion of tissues at the apex and base and because it best accounts for the spectrum of prostate contours [4, 7, 8, 14, 15]. In this assessment, RCAD over- or underestimated individual prostate volumes in only 4 cases (2.6%) (Figs. 5-8). These individual case errors are unapparent in statistics as they are balanced out when evaluating for group mean or median in large numbers of patients. That explains why these central values are similar for RCAD and ACAD in ANOVA statistics. Our experience agrees with that of Benxinque, et al.[10], that for any individual patient, automatic unadjusted segmentation must be monitored and manually corrected for extreme misregistration of boundaries.

**Fig. 5.**
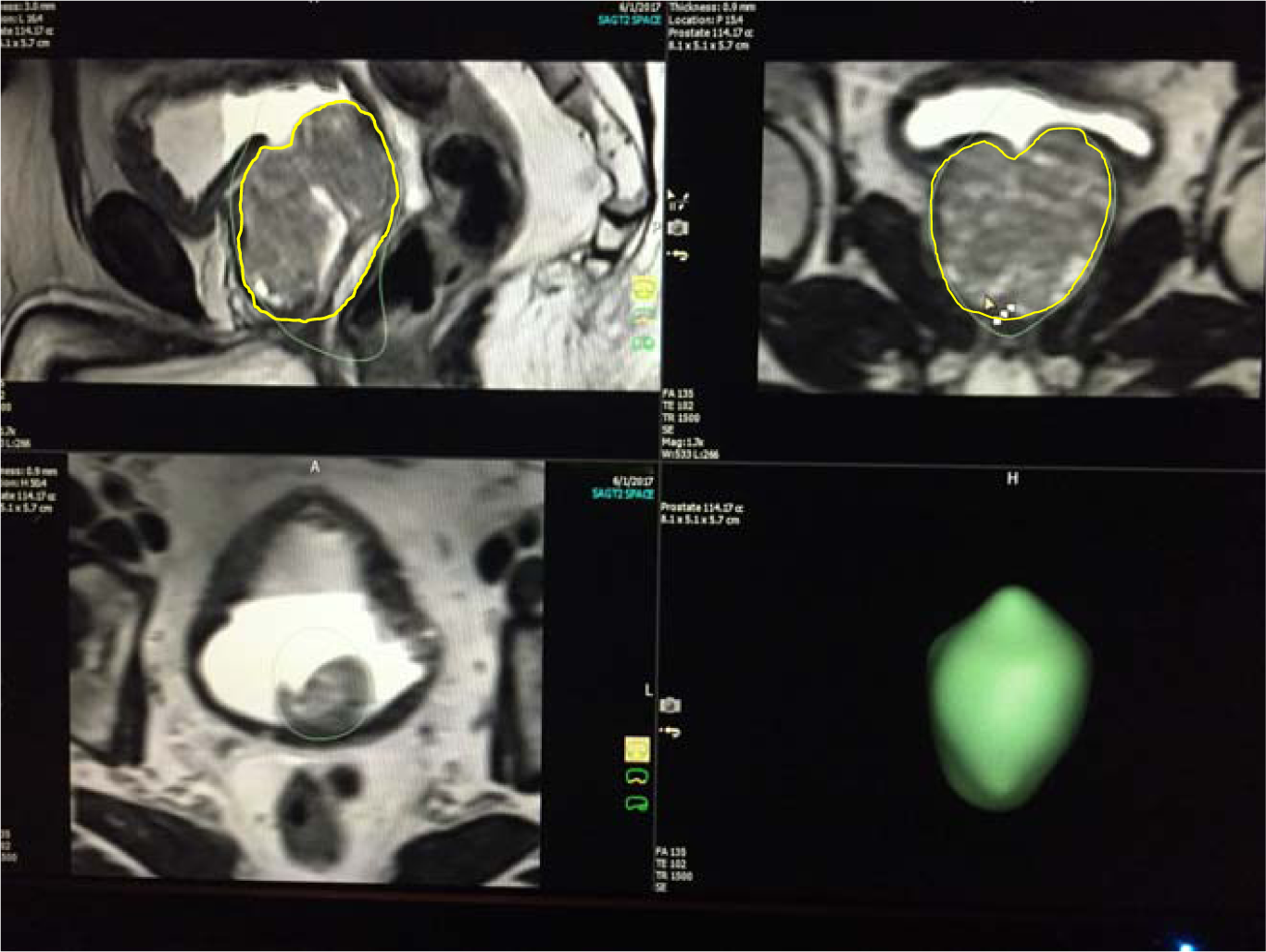
Unadjusted automatic DynaCAD^®^ over measurement error (green lines) indicating 114 cc total prostatic volume. Adjusted DynaCAD^®^ measurement (yellow lines) was 94 cc

**Fig. 6.**
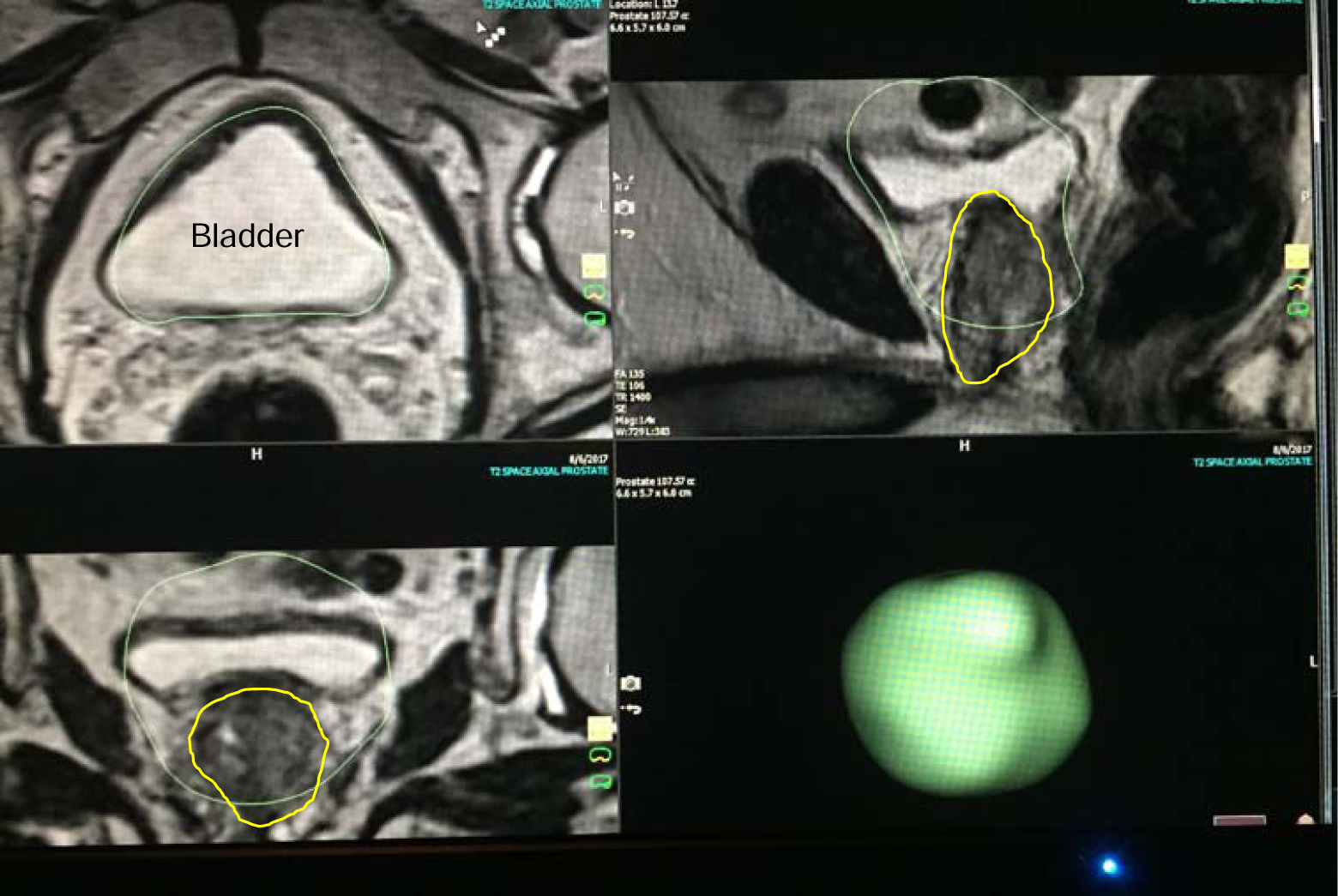
Unadjusted automatic DynaCAD^®^ over measurement error (green lines) indicating 108 cc total prostatic volume Adjusted DynaCAD^®^ (yellow lines) measurement was 28.5cc

**Fig. 7.**
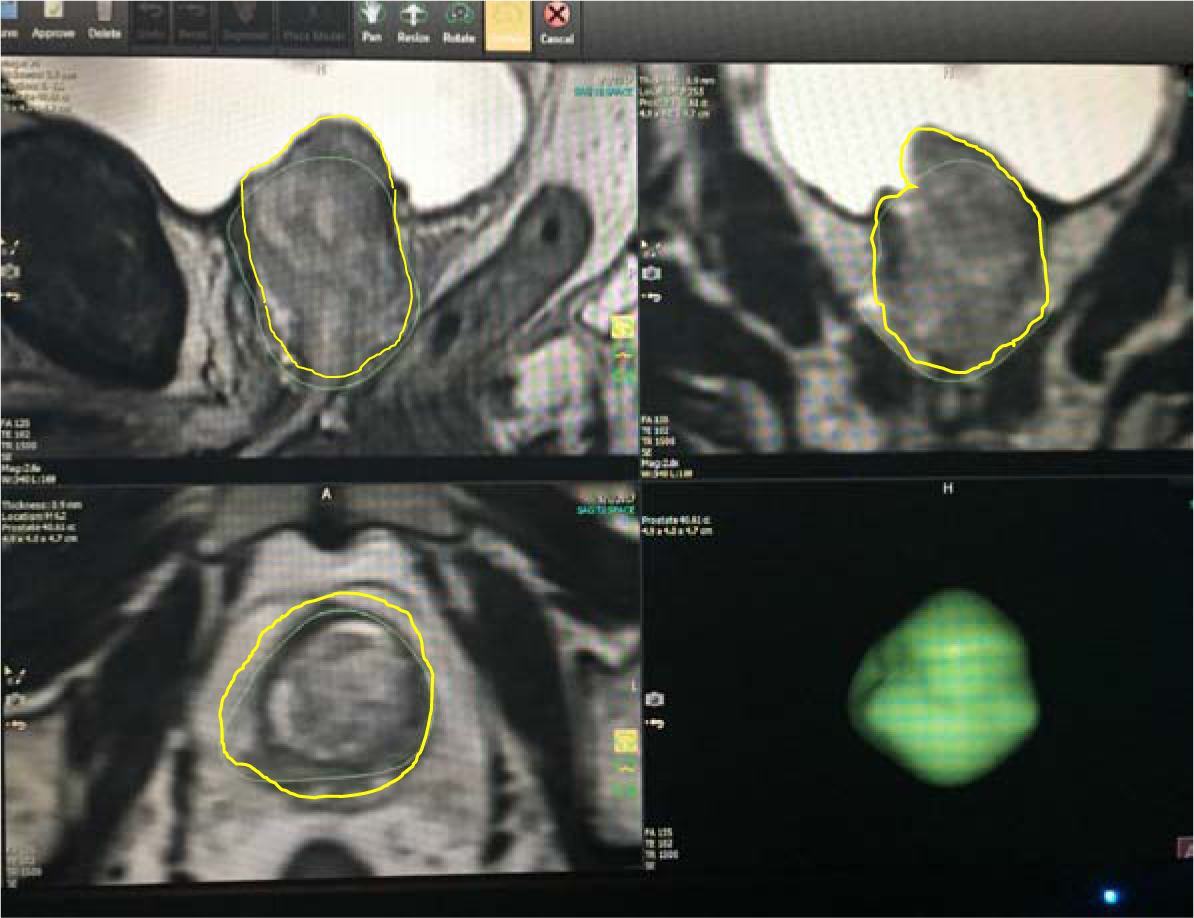
Unadjusted automatic DynaCAD^®^ under measurement error (green line) indicating 41 cc prostatic volume. Adjusted volume (yellow lines) measured 42.33

**Fig. 8.**
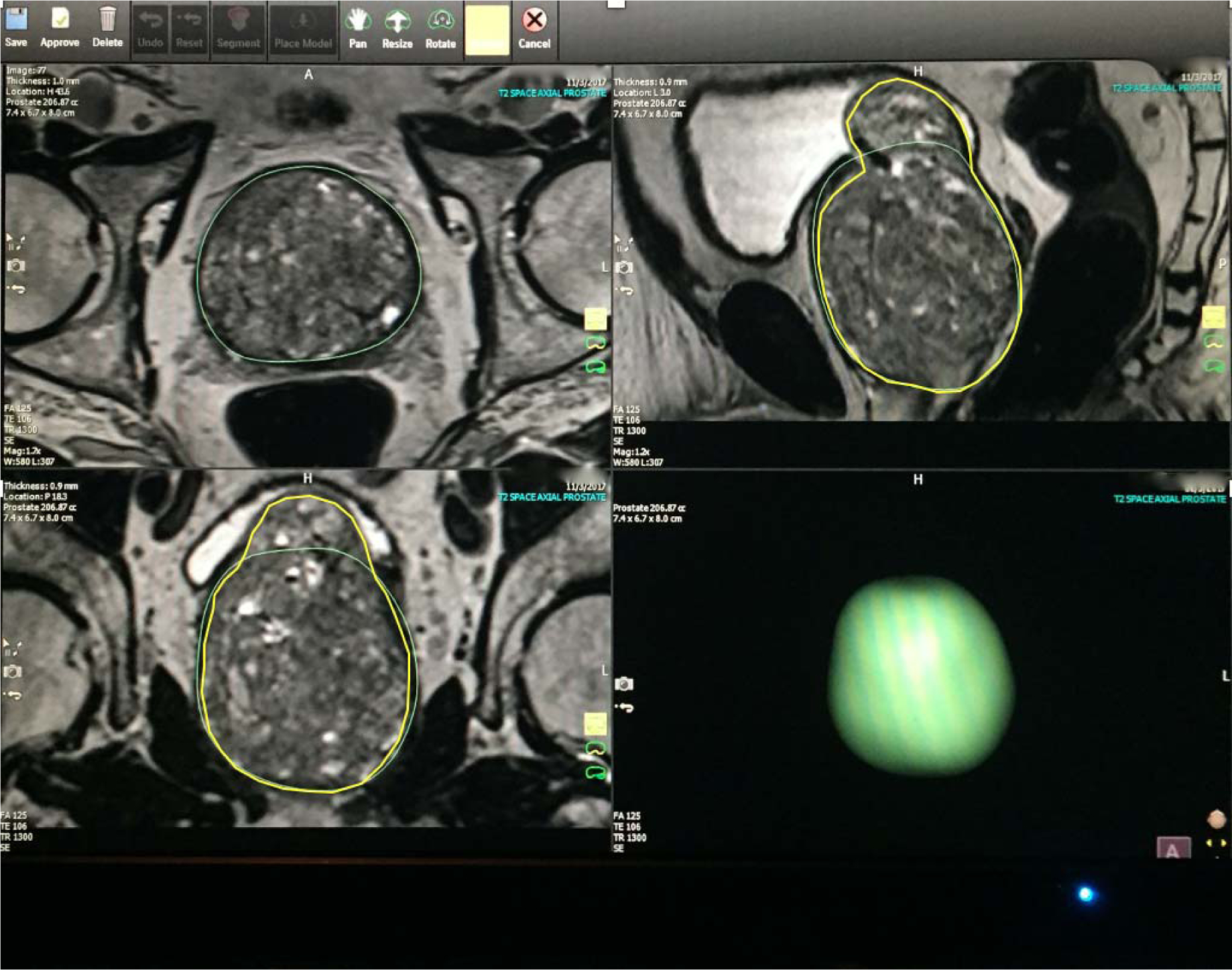
Unadjusted automatic DynaCAD^®^ under measurement error (green lines) indicating 207 cc. Adjusted DynaCAD^®^ measurement (yellow lines) was 282 cc

There are many issues using post-surgical or cadaver prostate specimens as the “ground truth” for prostate volume when comparing measurements utilizing ellipsoid or even planigraphic methods. Due to significant differences in the way the specimens were processed in these experiments, no true standard can be asssumed. A more robust discussion of this topic has been previously reported [3,11]. We should be more interested in measurements made *in situ* in the living blood -perfused patient for guidance in medical or surgical management. We are in agreement with Eri, LM, et al [14] and Christie, et al.[3] that it makes better sense to consider prostate MRI as the “gold standard” for reliable measurement of total prostate volume going forward in the future.

TRUS precision and accuracy data are available for ellipsoid and planigraphic volumetrics [1,16]. There are few studies examining precision and accuracy with MRI segmentation [9,10,17]. Turkbey, et al. [8] with non-commercially available software compared accuracy of fully automated segmentation, manual segmentation and ellipsoid volumetrics using post-operative prostate specimens as the “ground truth”. Pearson correlation coefficients were reported to be 0.90 for automated and manual segmentation and 0.86 for the ellipsoid. However, actual calculated volumes were significantly smaller compared with the specimens due to the retrospective nature of the experiment wherein parts or all of the specimen seminal vesicles, vas deferens were included in the weight along with periprostatic tissues. Therefore, though the imaging volumes were proportional to the specimen weights, they were not accurate using surgical specimens as the “gold” standard. Our results agree with that of Benxinque, et al. [10] who used DynaCAD^®^ (Invivo Corporation, Gainesville, FL) and also concluded that automated segmentation slightly overestimated prostate volume compared with manual segmentation and ellipsoid methods. Jeong,et al. [16] examined TRUS using the prolate ellipsoid model and planigraph-based MRI (Rapida version 2.8, Seoul Korea) compared with seminal vesicle free specimen volume in water. They demonstrated high accuracy for both MRI volume segmentation and TRUS. Karademir et al. [17] used ITK-SNAP software (Penn Image Computing and Science Laboratory) to obtain semiautomated prostate volumes and compared with specimens that included the seminal vesicles. They corrected that specimen weight by subtracting the estimated weight of 3.8 gm suggested to be the mean weight of the latter from the literature [18]. This resulted in high correlation between the MRI volume and specimen weight (accuracy) with a correlation coefficient (r) =0.94.

It is unclear how many examiners participated in volume acquisitions in the Turkbey study. The other studies were limited by the use of only one radiologist for the segmentation and ellipsoid volume measurements. Both were retrospective and specimens were not universally similarly handled prior to weighing and included extra-prostatic tissue. The latter group decided to use manually segmented measurement as the “gold standard” with which to compare other volume measures. Pasquier, et al. [19] using an atlas-based proprietary MRI algorithm and multiple readers, showed that automatic and manual results were comparable, but automated overestimated volume compared with manual. This is similar to our findings. None of the foregoing authors assessed inter-observer precision.

The main limitation of our study is that it is retrospective and all measurements were made by one observer, even though it was an examiner with experience and expertise. As stated earlier, this may reduce individual bias introduced by independent examiners. Our busy department clinical schedule could not accommodate for the time necessary to add 4 measurements by independent observers for each case.. Two or three examiners would have strengthened the study and allowed for inter-observer concordance analysis (precision). While unaware of baseline volume measurements in the medical record, bias introduced by the observer being aware of his own earlier measurements of volume cannot be excluded, especially in the case of the alternating ellipsoid measures done before segmentation.

In a 2019 white paper on PI-RADS^®^ v2.1, the American College of Radiology and European Society of Uroradiology advocate prostate volume measurement with “…manual or automated segmentation or calculated using (the) ellipsoid formula…” [20]. A study by Becker, et al. [21] using ITK-Snap software (itksnap.org) showed relatively high precision (0.75) of T2-weighted MRI manual segmentation measured by standard dice scores (DS=0.738), Hausdorff distance (HD=36.2), and volumetric similarity coefficient (VS=0.853). However, they also emphasize the labor intensiveness of their methods. There are no publications of DynaCAD^®^ performance utilizing these widely accepted measurements of precision performance, nor were they measured in this report.

Fully automatic planigraphic-based segmentation will eventually become the standard method for establishing total prostate volume. It is the only method that will completely eliminate interobserver reliability (precision) error. However, continuing insufficiencies exist for reliably finding prostatic outer boundaries limiting its use in today’s busy clinical environment.

At present, MRI T2-weighted manually adjusted segmentation (ACAD) is likely the most accurate method for determining prostatic total volume, because it it measures the summation of stacked axial segments conforming to the volume within the outer contours of the gland. This volume is calculated by mathematical algorithms, whereas ellipsoid methods rely on geometric models that “approximate” the prostatic contour. Our study suggests that may fulfill that function, but it must be monitored for gross misregistration of boundaries and manually adjusted. More research is needed to establish the repeatability (precision) of MRI-based planigraphic methods.

Optimal accuracy for total volume measurement has clinical importance. Segmentation applications such as DynaCAD^®^ may be applied in patients with suspected prostate cancer where accurate volume is incorporated into the prostate specific antigen density ratio (PSAD) urologists use to stratify risk for cancer in patients who have elevated PSA [22].

PSAD=Total PSA (ngm/mL) / prostate volume (ml^3^) [22,23]. PSAD is based on the concept that PSA production per ml^3^ of cancer tissue is increased by a factor of 10 compared to benign prostatic hyperplasia [24,25]. PSAD ≤ 0.12-0.15 favors benign disease [22,23]. However, others have not confirmed its value in the intermediate PSA range of 4.0 -10.0 ng/mL range [26]. PSAD in combination with PI-RADS has been shown to enhance predictive value for cancer detection[17,27,28]. PSAD is not only used in initial stratification of patients for biopsy and surgery, but also is being studied as a marker in the follow-up of patients in active surveillance [29,30]. The validity of MRI-based calculations of PSAD has been established 28,31]. Whether such a degree of incremental accuracy over ellipsoid methods will have a clinically significant impact on PSAD calculation or use is speculative. However, until speedy reliable fully automated MRI software is commercially available, ellipsoid formula methods remain quick and nearly as accurate for routine clinical work. ACAD will remain valuable in the research setting, but its main limiting factor is added user time for manual setting of prostate boundaries.

## Data Availability

Data is available from upon any reasonable request.

https://www.wasse001@umn.edu

## Notes

### Competing Interest Statement

The authors have declared no competing interest.

### Clinical Trial

NA

### Funding Statement

No external funding was used.

### Author Declarations

Institutional IRB and Ethics Committee (Ethsirb) 2018 approved.

